# Single-dose SARS-CoV-2 vaccine in a prospective cohort of COVID-19 patients

**DOI:** 10.1101/2021.05.25.21257797

**Authors:** Marit J. van Gils, Hugo D. van Willigen, Elke Wynberg, Alvin X. Han, Karlijn van der Straten, Anouk Verveen, Romy Lebbink, Maartje Dijkstra, Judith A. Burger, Melissa Oomen, Khadija Tejjani, Joey H. Bouhuijs, Brent Appelman, Ayesha H.A. Lavell, Meliawati Poniman, Tom G. Caniels, Ilja Bontjer, Lonneke A. van Vught, Alexander P.J. Vlaar, Jonne J. Sikkens, Marije K. Bomers, Rogier W. Sanders, Neeltje A. Kootstra, Colin Russell, Maria Prins, Godelieve J. de Bree, Menno D. de Jong, RECoVERED Study Group

## Abstract

**Background:** The urgent need for, but limited availability of, SARS-CoV-2 vaccines worldwide has led to widespread consideration of dose sparing strategies, particularly single vaccine dosing of individuals with prior SARS-CoV-2 infection.

**Methods:** We evaluated SARS-CoV-2 specific antibody responses following a single-dose of BNT162b2 (Pfizer-BioNTech) mRNA vaccine in 155 previously SARS-CoV-2-infected individuals participating in a population-based prospective cohort study of COVID-19 patients.

Participants varied widely in age, comorbidities, COVID-19 severity and time since infection, ranging from 1 to 15 months. Serum antibody titers were determined at time of vaccination and one week after vaccination. Responses were compared to those in SARS-CoV-2-naive health care workers after two BNT162b2 mRNA vaccine doses.

**Results:** Within one week of vaccination, IgG antibody levels to virus spike and RBD proteins increased 27 to 29-fold and neutralizing antibody titers increased 12-fold, exceeding titers of fully vaccinated SARS-CoV-2-naive controls (95% credible interval (CrI): 0.56 to 0.67 v. control 95% CrI: −0.16 to −0.02). Pre-vaccination neutralizing antibody titers had the largest positive mean effect size on titers following vaccination (95% CrI (0.16 to 0.45)). COVID-19 severity, the presence of comorbidities and the time interval between infection and vaccination had no discernible impact on vaccine response.

**Conclusion:** A single dose of BNT162b2 mRNA vaccine up to 15 months after SARS-CoV-2 infection provides neutralizing titers exceeding two vaccine doses in previously uninfected individuals. These findings support wide implementation of a single-dose mRNA vaccine strategy after prior SARS-CoV-2 infection.

## Introduction

The unprecedented rapid development and emergency use authorization of several vaccines against severe acute respiratory syndrome coronavirus 2 (SARS-CoV-2) allows for optimism in the global fight against the coronavirus disease 2019 (COVID-19) pandemic.^1^ However, in many regions, vaccination campaigns are hampered by limited supply or resources, hence vaccine sparing strategies are desirable. Making use of immunological memory after prior natural SARS-CoV-2 infection,^2,3^ single dosing represents one such strategy for vaccines requiring two doses for optimal efficacy. Indeed, a number of recent small studies in health care workers (HCW) have shown similar or higher antibody responses and higher vaccine efficacy to a single dose SARS-CoV-2 mRNA vaccine after prior infection, compared to two doses in SARS-CoV-2-naive individuals.^4–11^ However, these studies were performed in relatively young and healthy individuals and provided limited information on the possible influence of COVID-19 severity and the duration since infection on vaccine responses. To inform potential wide implementation of a single dose strategy following natural infection, we evaluated the titers and breadth of antibody responses to a single SARS-CoV-2 mRNA vaccine in an ongoing population-based prospective cohort study of COVID-19 patients, representing a range in age, the presence of comorbidities, COVID-19 severity and time since infection. Antibody responses were compared to those observed after two doses in a cohort of SARS-CoV-2 naive HCW.

## Methods

### Patients and study design

The current vaccine study was embedded in the RECoVERED project, an ongoing prospective cohort study of individuals with laboratory-confirmed SARS-CoV-2 infection in Amsterdam, the Netherlands. The RECoVERED cohort was initiated in May 2020 and as of April 2021 enrolled 328 participants, including both home-cared patients with mild infections and hospitalized patients with moderate to severe or critical illness. RECoVERED participants are followed at 1-3-month intervals from illness onset whereby biological specimens and questionnaires are collected at each follow-up visit to address RECoVERED’s primary objectives relating to immunology and long-term sequelae of COVID-19. Clinical severity groups were defined in line with the WHO COVID-19 disease severity criteria:^12^ Mild disease was defined as having a respiratory rate (RR) <20/min and oxygen saturation (SpO_2_) on room air >94% during acute illness; moderate disease as having a RR of 20-30/min, SpO_2_ 90-94% and/or receiving oxygen therapy; severe disease as having a RR>30/min or SpO_2_ <90%; and critical disease as requiring ICU admission.

Cohort participants, invited to receive vaccination according to the Dutch national vaccination campaign before 12 April 2021, were asked to participate in the present vaccine substudy. In addition, participants not yet prioritized for vaccination according to Dutch policy were asked to participate and receive a first dose of the BNT162b2 (Pfizer-BioNTech) mRNA vaccine in the week of April 12 2021, made available for our research aim by the Dutch Ministry of Health, Welfare and Sport. Participants with pregnancy, vaccine-related allergic reactions or laboratory-confirmed infections within 4 weeks of expected vaccination were excluded as per national guidelines. Serum samples for determination of antibody levels were collected and participants completed questionnaires on the presence and severity of symptoms and vaccine-related adverse effects within seven days pre-vaccination and within one week post-vaccination.

Vaccinated HCW without longitudinal serological evidence of prior SARS-CoV-2 infection, participating in a HCW cohort study at the Amsterdam University Medical Centers (S3 study), served as a control group.^13^ In this cohort, antibody responses were measured four weeks after the second dose of the BNT162b2 mRNA vaccine. The RECoVERED study, including the vaccine substudy, and the S3 study were approved by the medical ethical review board of the Amsterdam University Medical Centers (NL73759.018.20 and NL73478.029.20, respectively). All participants provided written informed consent.

### SARS-CoV-2 binding IgG antibody levels

Levels of Immunoglobulin G (IgG) binding to SARS-CoV-2 receptor-binding domain (RBD), nucleocapsid (N) and spike (S) proteins of wild type virus (Wu-1) and variants of concern (B.1.1.7, B.1.351 and P.1), as well as to control proteins tetanus toxoid, respiratory syncytial virus glycoprotein (RSV-G) and influenza A/H1N1pdm09 virus HA protein, were determined using a custom luminex assay as described previously.^14^ In short, proteins were produced in HEK293F cells (Invitrogen) and purified from the cell culture supernatant using affinity chromatography with NiNTA agarose beads (Qiagen). Proteins were covalently coupled to luminex magplex beads using a two-step carbodiimide reaction. Beads were incubated overnight with 1:100,000 diluted serum followed by detection with goat-anti-human IgG-PE (Southern Biotech) on a Magpix (Luminex) as the mean fluorescent intensity (MFI).

### Pseudovirus neutralization assay

Pseudovirus neutralization assay was performed as previously described (Brouwer et al. Cell 2021). Briefly, HEK293T/ACE2 cells^15^ were seeded in poly-L-lysine pre-coated 96-well plates. The next day, heat-inactivated 1:100 diluted sera were 3-fold serially diluted and mixed in a 1:1 ratio with pseudovirus Wu-1 D614G (WT)^15^ or B.1.1.7, B.1.351 and P.1 pseudovirus. After 1-hour incubation at 37°C the mixtures were added to the cells and incubated for 48 hours at 37°C. The luciferase activity in cell lysates was measured using the Nano-Glo Luciferase Assay System (Promega) and GloMax system (Turner BioSystems). The 50% inhibitory dilution (ID_50_) titers were determined as the serum dilution at which infectivity was inhibited by 50% using a non-linear regression curve fit (GraphPad Prism software version 8.3).

### Statistical analysis

A Bayesian hierarchical generalization of the one-way ANOVA model was used to compare control, pre- and post-vaccination neutralizing and IgG antibody binding titers. Differences between groups are reported as differences in effect sizes. A difference in effect size is non-trivial if it is non-zero, and substantial if greater/lower than 1/-1. This model was also used to estimate the individual effect size of age group, clinical severity, and time since symptom onset group, on the observed vaccine responses.

Furthermore, a Bayesian multilevel model that partially pooled effect size estimates across all study participants was used to estimate the effect size of the predictor variables individually and in combination on post-vaccination serum neutralization levels (Table S1). We investigated if, and the degree to which, participants’ age, sex, presence of comorbidities (i.e. history of cancer, cardiovascular disease, chronic respiratory disease, diabetes mellitus and obesity, separately), COVID-19 severity, time since COVID-19 symptom onset, pre-vaccination neutralization titers, and infection by a B.1.1.7 lineage variant were correlated with vaccine response. Condition indices were computed to ensure that there was no collinearity among the predictor variables (i.e. condition index <10). A distribution of normalized effect sizes (analogous to regression coefficients) was estimated for each predictor variable as a measure of their relative contributions to vaccine response. Similar to the Bayesian ANOVA model, an effect size is non-trivial if it is non-zero, and substantial if greater/lower than 1.

All Bayesian models were fitted using Markov Chain Monte Carlo (MCMC) with pymc3,^16^ implementing a no-u-turn sampler. Four MCMC chains were run with at least 4000 burn-in steps and 2000 saved posterior samples. Convergence for all parameters were verified by checking trace plots, ensuring their 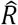 values were <1.05 with sufficient effective sample size (>200).

## Results

### Study population

A total of 155 participants of the RECoVERED cohort received a single-dose of the BNT162b2 mRNA vaccine after a median of nine months following SARS-CoV-2 infection (interquartile range (IQR) 5-12 months). The median age of participants was 55 years (IQR 33-61), 37% were female, 44% had one or more comorbidities and their SARS-CoV-2 infections were classified as mild, moderate, or severe/critical COVID-19 in 33%, 45% and 22% of participants, respectively (Table 1). Overall, the vaccine was well tolerated with only mild and self-limiting adverse events (Table S2). A total of 130 of the 155 participants (84%) reported one or more side effects within 48 hours of vaccination, with pain at the injection-site (84%) and fatigue (48%) reported most frequently. The control group consisted of 49 healthy HCW (62% female, median age 44 years (IQR 33-53)) without evidence of previous SARS-CoV-2 infection who received two doses of the BNT162b2 mRNA vaccine.

**Table 1:** Characteristics of the study participants.

|  | No. of participants (%) |
| --- | --- |
| Total | 155 |
| Age (median 55y; range (22 - 88y)) |  |
| <= 45 y | 55 (35.5%) |
| 46-65 y | 75 (48.4%) |
| > 65 y | 25 (16.1%) |
| Sex |  |
| Female | 58 (37.4%) |
| Male | 97 (62.6%) |
| Disease severity |  |
| Mild | 51 (32.9%) |
| Moderate | 70 (45.1%) |
| Severe | 15 (9.7%) |
| Critical | 19 (12.3%) |
| Time between symptom onset and vaccination<br>(median 9 months; range (1 - 15 months)) |  |
| <= 6 months | 60 (38.7%) |
| 7-12 months | 60 (38.7%) |
| >12 months | 35 (22.6%) |
| Comorbidities |  |
| Cancer | 8 (5.2%) |
| Cardiovascular disease | 29 (18.7%) |
| Chronic respiratory disease | 23 (14.8%) |
| Diabetes Mellitus | 14 (9.0%) |
| Obesity | 28 (18.1%) |

### SARS-CoV-2 IgG antibody responses

Prior to vaccination, levels of IgG antibodies binding to S, RBD and N proteins exhibited a wide range, with overall higher levels observed in participants with previous severe/critical COVID-19 (Figure 1A, S1A). Sharp increases in anti-S and anti-RBD IgG were observed 1 week after vaccination (median fold increase 29.4 (IQR 9.1-92.1) and 27.6 (IQR 10.3-70.8), respectively). Using a Bayesian ANOVA model, we found substantial differences between pre- and post-vaccination for anti-S and anti-RBD IgG levels (95% credible interval (CrI) of difference in effect size: 1.30 to 1.55 for anti-S and 1.29 to 1.57 for anti-RBD; Figure S1B-C; Table S3). Achieved levels were similar or higher to those observed four weeks after two vaccinations in the SARS-CoV-2-naive HCW control group (Figure S1B; Table S3). When looking at anti-S IgG to three variants of concern (VOC; B.1.1.7, B.1.351 and P.1), levels were comparable to wild-type (WT) S protein both pre- and post-vaccination, with discernible increases for all VOC S proteins after vaccination (Figures 1B, S2A; Table S3).

**Figure 1:**
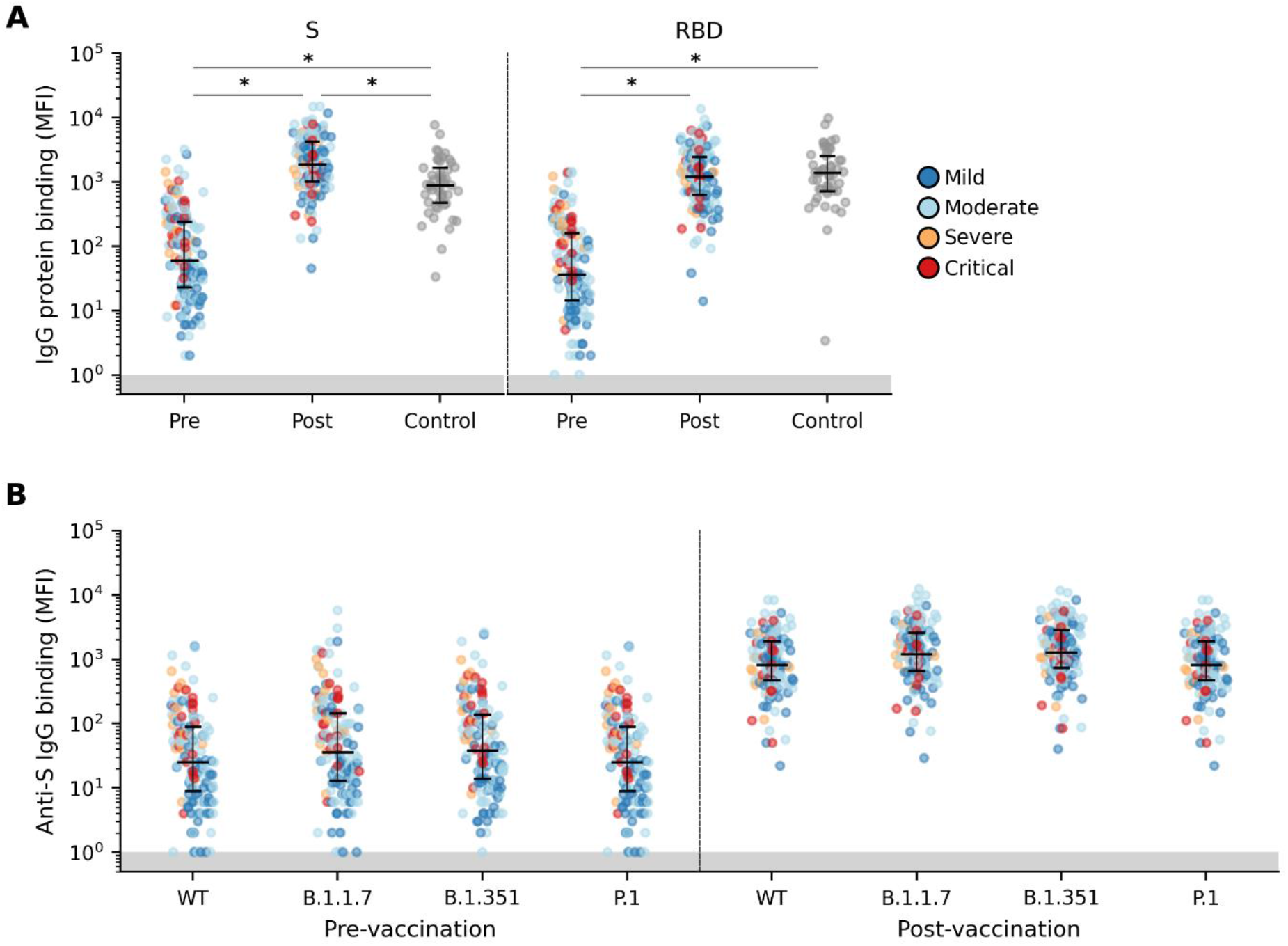
Anti-SARS-CoV-2 IgG responses after mRNA vaccination. (**A**) Pre- (n=153 participants), post-vaccination (n=155) and HCW control (n=49) distributions of serum IgG levels to SARS-CoV-2 S and RBD protein. (**B**) Pre- and post-vaccination distributions of anti-S IgG levels to WT and VOC lineages B.1.1.7, B.1.351 and P.1. Each point represents one participant colored by COVID-19 severity. Median with interquartile range is depicted. *; distributions with non-overlapping 95% CI of group mean effect size estimated using a Bayesian ANOVA model (Table S3). Area of binding values below detection limit are shaded in gray.

### SARS-CoV-2 neutralizing antibody responses

Neutralizing antibodies were detected (ID_50_ >100) prior to vaccination in 134 of 155 (86%) participants, correlating with COVID-19 severity and time since infection. Similar to SARS-CoV-2-specific IgG, neutralization titers increased sharply after vaccination (median fold increase 12.5 (IQR 5.2-39.6)), achieving higher titers than observed in the control group (Figure 2A, S1A-B, Table S4). Bayesian multilevel regression analysis showed that anti-S IgG (95% CrI: 0.29, 0.88) correlated more strongly to neutralization levels than anti-RBD IgG (CrI: 0.08, 0.66) before vaccination. However, after vaccination anti-RBD IgG correlated strongly with neutralization (95% CrI: 0.55, 1.41), while no discernable effects of anti-S IgG were observed (CrI: −0.55, 0.28); Figure S3). This indicates that neutralization is dominated by IgG binding to RBD after vaccination.

**Figure 2:**
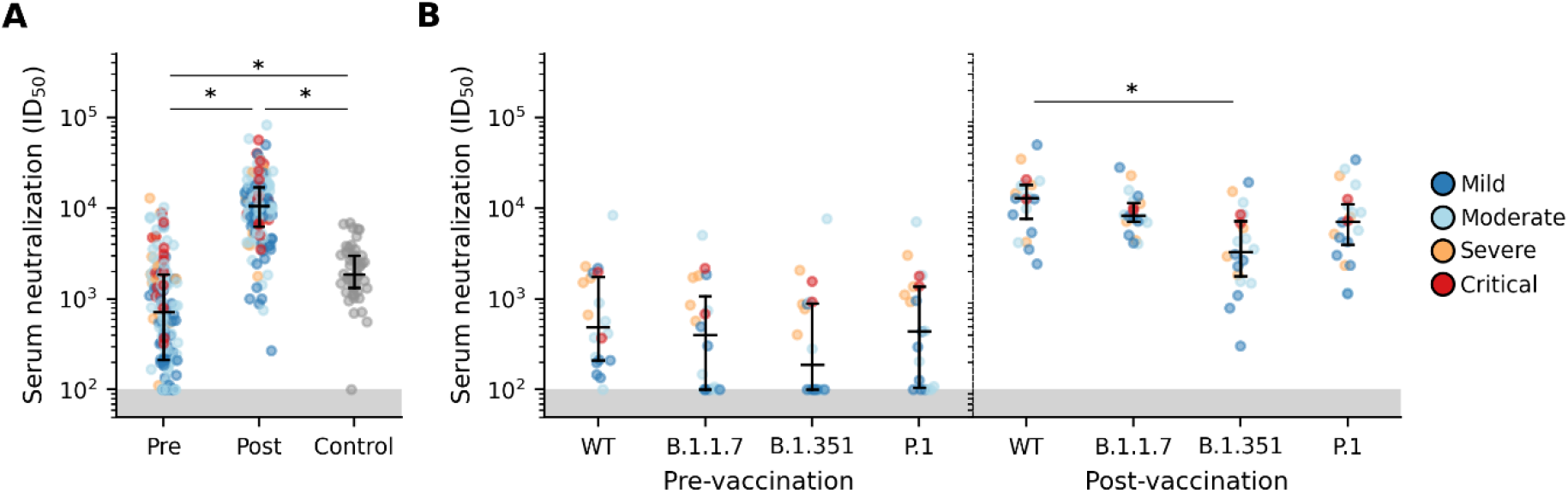
Serum neutralization of SARS-CoV-2 pseudovirus after mRNA vaccination. (**A**) Pre- (n=153 participants), post-vaccination (n=155) and HCW control (n=49) distributions of SARS-CoV-2 pseudovirus neutralization. (**B**) Pre- and post-vaccination serum neutralization distributions of WT and VOC lineages B.1.1.7, B.1.351 and P.1 using sera from a random selection of 20 participants. Each point represents one participant colored by COVID-19 severity. Median with interquartile range is depicted. *; distributions with non-overlapping 95% CI of group mean effect size estimated using a Bayesian ANOVA model (Table S4). Area of neutralization titers below detection limit are shaded in gray.

Neutralization of VOC was evaluated in a random selection of 20 participants with detectable WT neutralization titers. While 11 of these 20 participants had undetectable neutralizing activity against one or more VOC before vaccination, neutralization titers rose sharply and were measurable in all participants after vaccination to all three VOC (median fold increase 29.8 (IQR 7.2-70.4; 95% CrI: 1.05, 1..64), 9.1 (IQR 6.4-21.2; 95% CrI: 0.67, 1.37) and 10.5 (IQR, 5.6-36.8; 95% CrI: 0.78, 1.46) for B.1.1.7, B.1.351 and P.1, respectively; Figure 2B, S2B; Table S4). The differences between WT and VOC neutralization titers were trivial, except for B.1.351 post-vaccination (95% CrI: −0.90, −0.08), with the ratio between WT and VOC neutralization diminished after vaccination (Figure S2C; Table S4).

### Predictors of vaccine response

We used a Bayesian multilevel regression model to estimate the effect size of variables potentially affecting post-vaccination neutralization levels in 139 participants from whom complete metadata were available (Table S5). Pre-vaccination neutralization levels showed the largest positive mean effect with clear posterior support of non-trivial effect size (95% CrI: 0.16, 0.45). In addition, age (95% CrI: −0.29, −0.05) and sex (CrI: 0.04, 0.32) exhibited modest non-trivial effects (Figure 3A). Further analyses using the Bayesian ANOVA model showed that mean post-vaccination neutralization levels were discernible for sex (95% CrI: male: −0.15, 0.00; female: 0.00, 0.15) but not between different age groups (95% CrI: ≤45y: −0.05, 0.16; 46-65y: −0.08, 0.12; >65y: −0.24, 0.06; Figure S4A). This indicates that, while younger individuals were expected to achieve higher neutralizing responses, neutralization levels overlapped widely between different age groups. Post-vaccination neutralization titers and fold increases after vaccination were similar across COVID-19 severity groups and were not affected by comorbidities. The time period between SARS-CoV-2 infection and vaccination also did not affect post-vaccination neutralization levels, nor were discernible effects observed in fold increases of neutralization titers when comparing different intervals between infection and vaccination, i.e. ≤6 months, 7-12 months and >12 months (Figure 3B).

**Figure 3:**
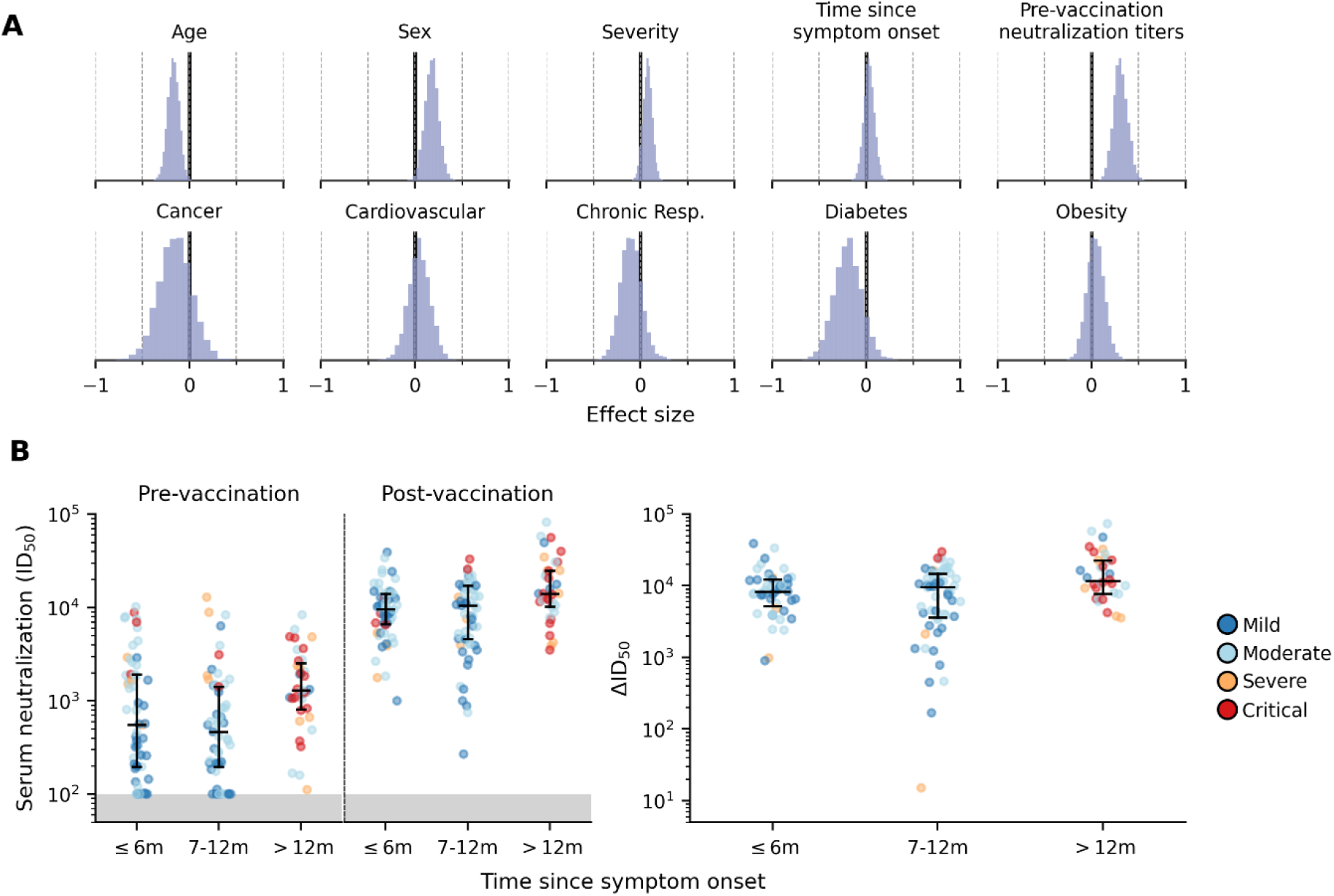
Predictors of vaccine response. (**A**) Joint contributions of participant and clinical factors on post-vaccination serum neutralization titers. The mean effects across study participants were estimated using a Bayesian multilevel model. All continuous predictors were mean-centered and scaled such that effect sizes shown can be compared on a common scale. (**B**) Distributions of pre- and post-vaccination (left panel) as well as fold change (ΔID_50_; right panel) in serum neutralization titers of study participants stratified according to time since illness onset. Each point represents one participant colored by COVID-19 severity. Area of neutralization titers below detection limit are shaded in gray.

Seven participants were infected with a B.1.1.7 lineage variant. While the binding and neutralizing antibodies responses for these individuals fell within the range of those observed in participants infected with non-VOC variants (Figure S4B), the small number of B.1.1.7 infected individuals prevented reliable assessment of any statistically meaningful differences. Importantly, inferences on effect sizes of possible factors affecting post-vaccination neutralization titers were not affected by inclusion of B.1.1.7 infected participants (Table S6).

## Discussion

This study demonstrates that higher levels of neutralizing antibodies are achieved within a week after a single dose of a SARS-CoV-2 mRNA vaccine in previously infected individuals compared to those observed in fully vaccinated SARS-CoV-2 naive HCW, irrespective of time since infection. This implies that a single-dose in prior-infected individuals administered up to more than one year after SARS-CoV-2 infection provides antibody responses associated with the vaccine efficacy observed in the phase 3 study of the BNT162b2 mRNA vaccine.^17^ Similar favorable vaccine responses after natural infection have been reported but these studies were fairly small, restricted to relatively young and healthy HCW with mostly mild disease who were vaccinated up to nine months after infection.^4–11^ Our prospective COVID-19 cohort allowed extension of these findings to a broader population at risk and showed that these responses were not affected by the presence of underlying comorbidities, COVID-19 disease severity or timing of vaccination since infection. Hence, our study supports wide implementation of single dosing strategies for previously infected individuals.

The Bayesian multilevel regression model showed that pre-vaccination neutralization titers were associated with higher neutralization titers after vaccination, independent of disease severity or time since infection. This may suggest that pre-existing antibodies potentially augment immune responses perhaps through the formation of immune complexes by antibodies binding the vaccine antigen.^18,19^ In keeping with other studies,^6,20,21^ age and male sex were inversely correlated with vaccine responses. Even though variation in the neutralization titers was large and effect sizes overlapped between the age groups, younger individuals, especially females, are expected to have higher neutralization responses, which have also been observed with other vaccines (e.g. influenza).^22,23^

The emergence of SARS-CoV-2 variants may pose risks of infection despite immunity induced by natural infection and vaccination. Indeed, emerging observations indicate substantial reductions of vaccine-induced antibodies in binding and neutralization capacity against several VOCs, including B.1.351 and P.1.^9,24–26^ After single vaccination, we found sharp increases of IgG binding and neutralization levels for the three VOC. However, neutralization titers for the B.1.351 variant lagged behind those for wild type and other VOCs. Overall, these results suggest that neutralization breadth was not improved after vaccination, most likely because neutralization after vaccination is overwhelmingly dominated by RBD responses, which are shown to be more sensitive to the mutations in the VOC.^27^ Nevertheless, a degree of cross-neutralization of these three VOCs was observed in all participants already after a single dose in previously infected individuals.

There are several limitations of our study. As only symptomatic COVID-19 patients were followed we were unable to study vaccine responses after previous asymptomatic SARS-CoV-2 infection. However, an earlier study in HCW has observed no differences in antibody responses to a mRNA vaccine between individuals with prior asymptomatic and symptomatic SARS-CoV-2 infections.^10^ Furthermore, the SARS-CoV-2 naive HCW controls were not matched with the previously infected cohort participants for potentially relevant factors such as age, sex or the presence of comorbidities. However, given that antibody responses in the healthier and younger HCW controls were lower, combined with our finding that age is inversely correlated with antibody vaccine response, the observed difference in vaccine response might even have been more pronounced if controls were matched. Finally, participants with severe COVID-19 were overrepresented in the subgroup with >12 months interval between infection and vaccination, but fold increases in neutralization were very similar for all time interval subgroups, independent of disease severity.

Longer follow up will determine the longevity of the immune response and protective efficacy after single dosing in previously infected individuals. In addition, while similar antibody boost responses would be anticipated for other SARS-Co V-2 vaccines, this needs to be confirmed in future studies, especially for non-mRNA vaccines. In the meantime, the findings of this study support wide implementation of a single-dose mRNA vaccine strategy after prior SARS-CoV-2 infection to save vaccines and resources hence expediting vaccination uptake at community levels worldwide.

## Data Availability

Data supporting the findings in this manuscript are available from the corresponding author upon request.

## Acknowledgements

The authors wish to thank all RECoVERED and S3 study participants; Mathieu A. F. Claireaux of Amsterdam UMC, Amsterdam, the Netherlands for providing the control proteins; Gestur Vidarsson and Federica Linty of Sanquin, Amsterdam, The Netherlands for providing the SARS-CoV-2 Nucleocapsid protein; Mohamed Hoesein, Marco Hol, Melvin Angelovski and their team of the Public Health Service of Amsterdam for valuable logistical support of the vaccine study; and the Amsterdam UMC COVID-19 S3/HCW study group (D. van de Beek, M.C. Brouwer, D.T.P. Buis, N. Chekrouni, N. van Mourik, S.E. Olie, E.J.G. Peters, T.D.Y. Reijnders, M. Schinkel, M.A. Slim, W.J. Wiersinga).

## Declaration of interests

All authors declare no competing interests.

## Funding

This work was supported by the Netherlands Organization for Scientific Research (NWO) ZonMw (RECoVERED, grant agreement no. 10150062010002 to M.D.d.J., S3 study, grant agreement no. 10430022010023 to J.J.S. and Vici grant no. 91818627 to R.W.S.), by the Bill & Melinda Gates Foundation (grant no. INV-002022 and INV008818 to R.W.S. and INV-024617 to M.J.v.G.), by Amsterdam UMC through the AMC Fellowship (to M.J.v.G.) and the Corona Research Fund (to M.K.B.), by the European Research Council (no. 818353 to C.A.R.), by the European Union’s Horizon 2020 program (RECoVER, grant no. 101003589 to M.D.d.J), and by the Public Health Service of Amsterdam (Research & Development grant no. 21-14 to M.P.). The funders had no role in study design, data collection, data analysis, data interpretation or data reporting.

## Notes

### Competing Interest Statement

The authors have declared no competing interest.

### Author Declarations

The RECoVERED study, including the vaccine substudy, and the S3 study were approved by the medical ethical review board of the Amsterdam University Medical Centers (NL73759.018.20 and NL73478.029.20, respectively). All participants provided written informed consent.

