## Supplementary material for "Single-dose SARS-CoV-2 vaccine in a prospective cohort of COVID-19 patients": Table S

**This PDF file includes:**
Figure S1 to S4
Table S1 to S6
Supplementary Methods

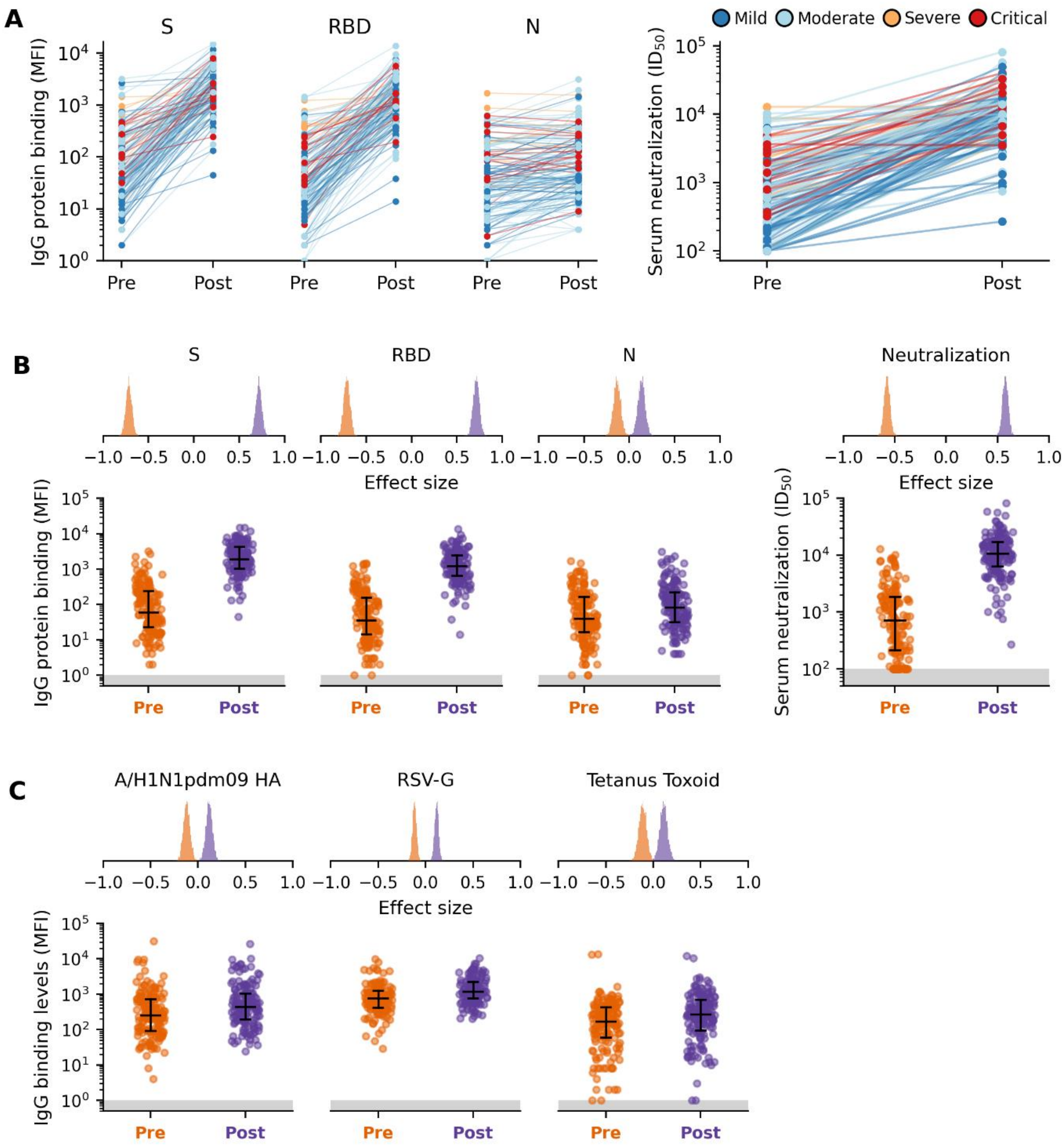


**Figure S1: Anti-SARS-CoV-2 antibody responses after mRNA vaccination**

(**A**) Paired pre- and post-vaccination IgG binding levels to S, RBD and N protein (left panel) and serum neutralization levels to WT SARS-CoV-2 (right panel). Each line between pre- and post-vaccination data points show the changes in binding levels for a study participant (colored by disease severity). (**B**) Pre- (orange) and post-vaccination (purple) distributions of IgG binding levels to S, RBD and N (left panel) and serum neutralization of SARS-CoV-2 (right panel). (**C**) Pre- and post-vaccination distributions of IgG binding levels to other human pathogens including influenza A/H1N1pdm09 haemagglutinin (HA) protein, respiratory syncytial virus glycoprotein (RSV-G) and Tetanus toxoid. The corresponding distribution of mean effect size estimates (Table S3) using a Bayesian ANOVA model is shown above each response distribution plot for (B) and (C).

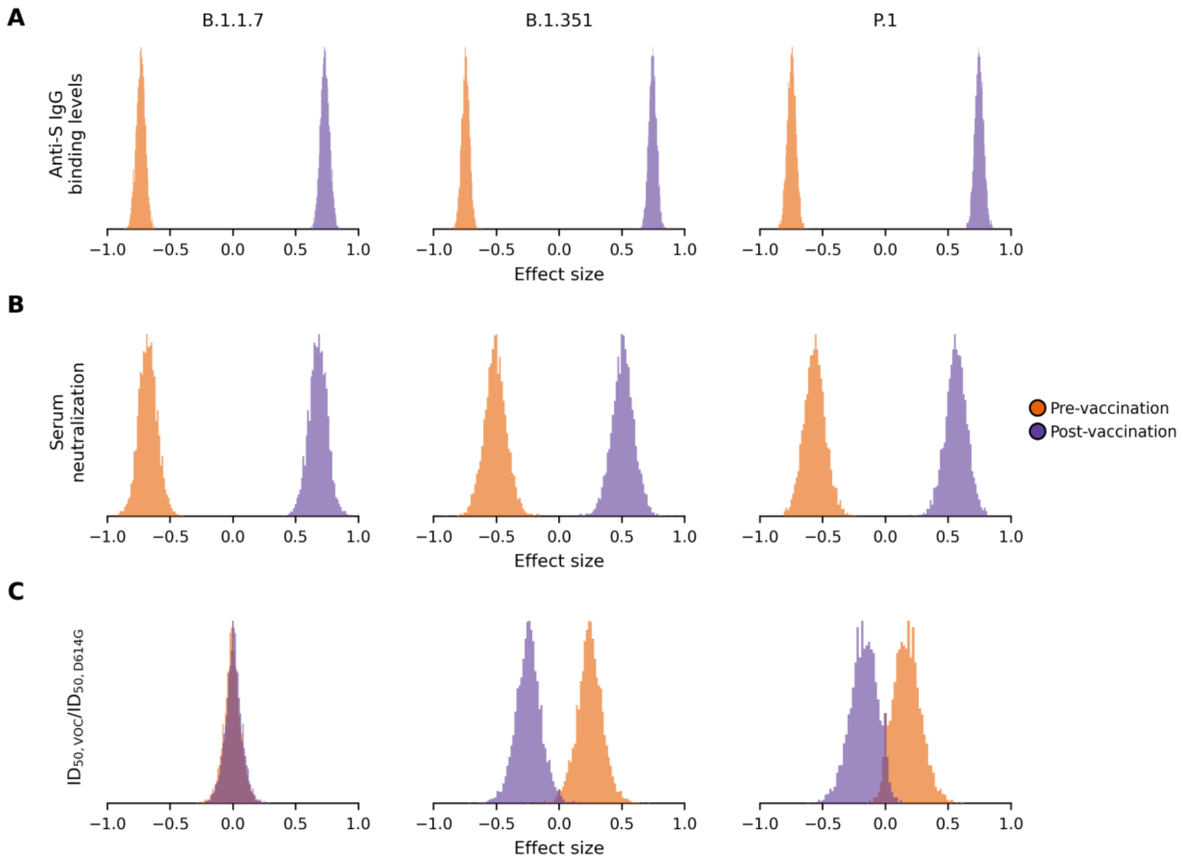


**Figure S2**: **Effect size estimates of pre- and post-vaccination antibody response after mRNA vaccination**

Pre- (orange) and post-vaccination (purple) distributions of mean effect size estimates (Table S4) using a Bayesian ANOVA model for (**A**) anti-S IgG binding, (**B**) serum neutralization levels and (**C**) ratio of neutralization against VOC to WT (D614G) for lineage B.1.1.7, B.1.351 and P.1.

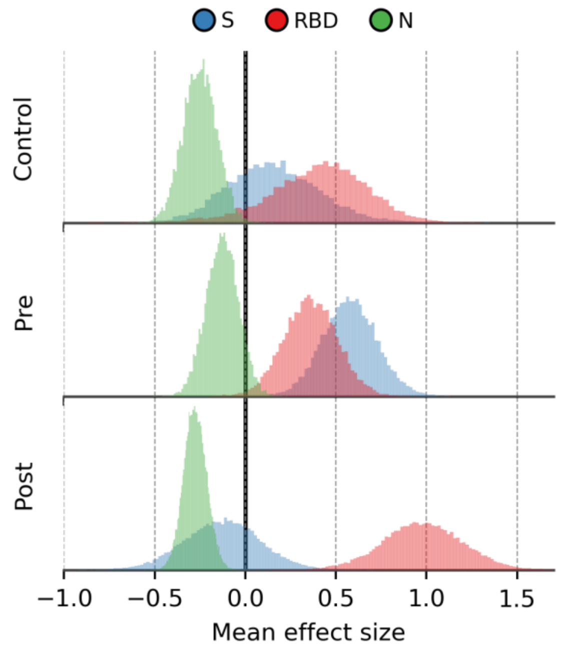


**Figure S3**: **Joint contributions of IgG binding to different SARS-CoV-2 antigens on control pre- and post-vaccination serum neutralization levels.**

The mean effects across study participants were estimated using a Bayesian multilevel model. All continuous predictors were mean-centered and scaled such that effect sizes shown can be compared on a common scale. S, spike protein; RBD, receptor binding domain protein; N, nucleocapsid protein

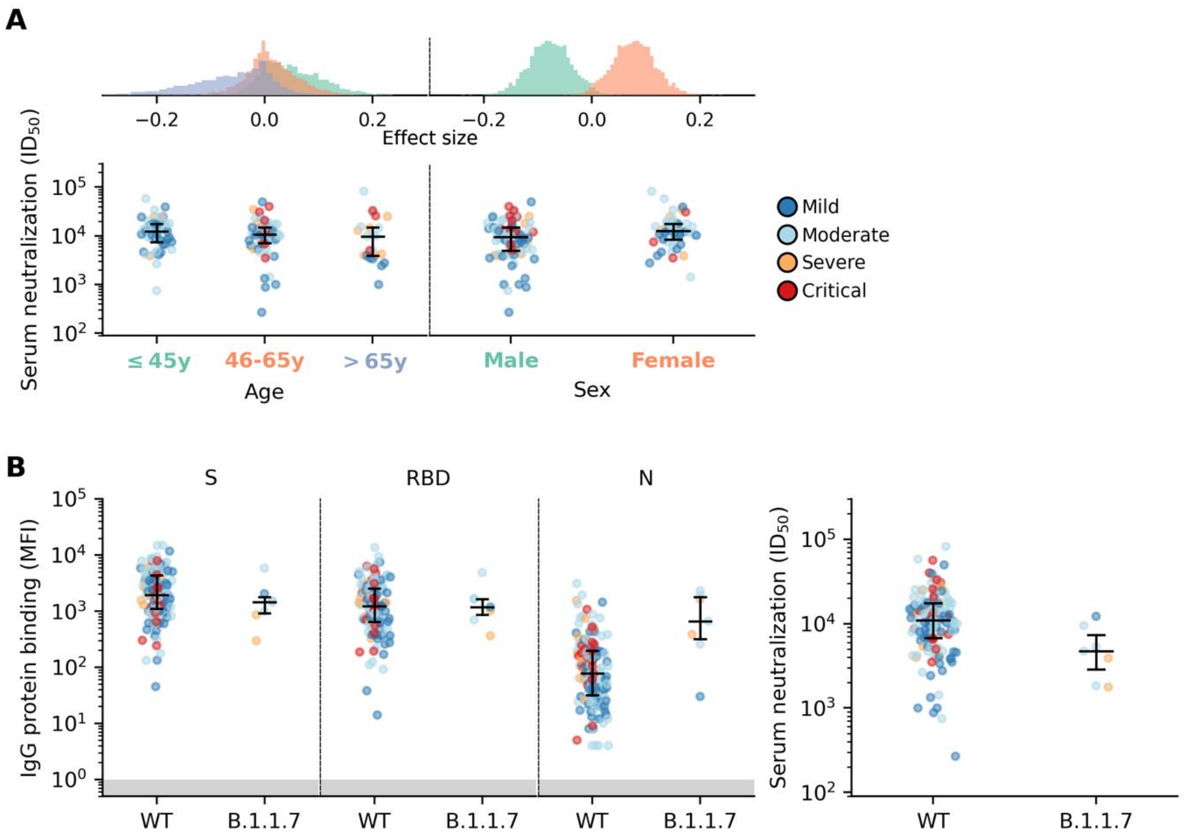


**Figure S4**: **Anti-SARS-CoV-2 antibody responses after mRNA vaccination stratified by age, sex and infection by lineage B.1.1.7.**

(**A**) Distributions of mean effect size estimates using a Bayesian ANOVA model (top) and post-vaccination serum neutralization levels (bottom) of study participants stratified according to age and sex. (**B**) Distributions of post-vaccination serum IgG binding and neutralization levels of study participants stratified according to the lineage of the infecting virus (wild-type (WT) or B.1.1.7).

**Table S1: Analyses performed using the Bayesian multilevel joint-contribution model**

| Analysis | Observed variable | Predictor variable |
|---|---|---|
| Correlating pre-vaccination neutralization titers to anti-SARS-CoV-2 IgG binding levels | Pre-vaccination neutralization titers | Pre-vaccination IgG binding levels to spike, RBD and nucleoprotein (all continuous) |
| Correlating post-vaccination neutralization titers to anti-SARS-CoV-2 IgG binding levels | Post-vaccination neutralization titers | Post-vaccination IgG binding levels to spike, RBD and nucleoprotein (all continuous) |
| Correlating post-vaccination neutralization titers to participant-specific meta variables | Post-vaccination neutralization titers | Age (categorical; ≥45years, 46-65years, >65years)<br><br>Sex (binary, male or female)<br><br>Severity (categorical; mild, moderate, severe / critical)<br><br>Time since symptom onset (categorical; ≥6 months, 7-12 months, >12 months)<br><br>Pre-vaccination neutralization titers (continuous)<br><br>Comorbidities (i.e. cancer, cardiovascular diseases, chronic respiratory disease, diabetes mellitus, obesity; binary, presence or absence)<br><br>Participant infected with the B.1.1.7 lineage variant (i.e. binary, presence or absence) |

**Table S2: Symptoms post-vaccination**

| Side effects | No. of participants (%) |
|---|---|
| Local | |
| Injection site pain | 130 (83.9%) |
| Injection site redness | 8 (5.2%) |
| Injection site swollen | 19 (12.3%) |
| Systemic | |
| Fatigue | 75 (48.4%) |
| Fever | 31 (20.0%) |
| Headache | 55 (35.5%) |
| Cold shivers | 34 (21.9%) |
| Myalgia | 41 (26.5%) |
| Arthralgia | 13 (8.4%) |
| Nausea/vomiting | 10 (6.5%) |
| Diarrhoea | 6 (3.9%) |

**Table S3: Bayesian ANOVA regression results: IgG binding**

| IgG binding levels (MFI) | | | | Bayesian ANOVA results | | | | |
|---|---|---|---|---|---|---|---|---|
| | | Interquartile range | | | 95% credible interval | | | |
| Variable | Median | Lower | Upper | Variable | Lower | Upper | ESS | $\hat{R}$ |
| Anti-S IgG against WT SARS-CoV-2 | | | | | | | | |
| Control | 887.3 | 479.9 | 1649.6 | Control | 0.14 | 0.33 | 962 | 1.00 |
| Pre | 60.0 | 23.0 | 242.0 | Pre | -0.91 | -0.74 | 956 | 1.00 |
| Post | 1888.0 | 1028.0 | 4233.0 | Post | 0.52 | 0.67 | 954 | 1.00 |
| | | | | Diff (Post-Control) | 0.23 | 0.51 | | |
| | | | | Diff (Pre-Control) | -1.24 | -0.91 | | |
| Fold change (Post-Pre/Pre) | 29.4 | 9.1 | 92.1 | Diff (Post-Pre) | **1.30** | **1.55** | | |
| Anti-RBD IgG against WT SARS-CoV-2 | | | | | | | | |
| Control | 1406.4 | 725.1 | 2543.1 | Control | 0.40 | 0.59 | 1041 | 1.01 |
| Pre | 36.0 | 14.5 | 158.0 | Pre | -1.05 | -0.87 | 1040 | 1.01 |
| Post | 1217.0 | 643.0 | 2457.0 | Post | 0.40 | 0.55 | 1035 | 1.01 |
| | | | | Diff (Post-Control) | -0.17 | 0.12 | | |
| | | | | Diff (Pre-Control) | -1.64 | -1.29 | | |
| Fold change (Post-Pre/Pre) | 27.6 | 10.3 | 70.8 | Diff (Post-Pre) | **1.29** | **1.57** | | |
| Anti-N IgG against WT SARS-CoV-2 | | | | | | | | |
| Control | 5.5 | 3.1 | 9.4 | Control | -0.78 | -0.62 | 799 | 1.01 |
| Pre | 40.0 | 16.5 | 164.0 | Pre | 0.12 | 0.30 | 788 | 1.01 |
| Post | 83.0 | 32.0 | 224.0 | Post | 0.41 | 0.57 | 791 | 1.01 |
| | | | | Diff (Post-Control) | 1.05 | 1.32 | | |
| | | | | Diff (Pre-Control) | 0.76 | 1.06 | | |
| Fold change (Post-Pre/Pre) | 0.9 | 0.0 | 2.5 | Diff (Post-Pre) | **0.14** | **0.43** | | |
| Tetanus toxoid | | | | | | | | |
| Pre | 168.0 | 60.0 | 435.0 | Pre | -0.19 | -0.03 | 763 | 1.01 |
| Post | 272.0 | 95.0 | 703.0 | Post | 0.03 | 0.19 | 757 | 1.01 |
| Fold change (Post-Pre/Pre) | 0.6 | -0.1 | 2.7 | Diff (Post-Pre) | **0.05** | **0.37** | | |
| RSV G | | | | | | | | |
| Pre | 771.0 | 421.0 | 1255.5 | Pre | -0.16 | -0.07 | 771 | 1.01 |
| Post | 1192.0 | 776.0 | 2252.0 | Post | 0.07 | 0.16 | 765 | 1.01 |
| Fold change (Post-Pre/Pre) | 0.6 | -0.1 | 2.4 | Diff (Post-Pre) | **0.15** | **0.31** | | |
| A/H1N1pdm09 HA | | | | | | | | |
| Pre | 252.0 | 92.0 | 730.0 | Pre | -0.19 | -0.05 | 982 | 1.00 |
| Post | 446.0 | 198.0 | 1052.0 | Post | 0.05 | 0.19 | 990 | 1.00 |
| Fold change (Post-Pre/Pre) | 0.7 | -0.1 | 2.6 | Diff (Post-Pre) | **0.10** | **0.37** | | |

| | | | | | | | | |
|---|---|---|---|---|---|---|---|---|
| Anti-S IgG against B.1.1.7 | | | | | | | | |
| Pre | 36.0 | 13.0 | 147.0 | Pre | -0.81 | -0.67 | 1227 | 1.00 |
| Post | 1212.0 | 654.0 | 2582.0 | Post | 0.67 | 0.81 | 1251 | 1.00 |
| Fold change (Post-Pre/Pre) | 31.2 | 11.0 | 93.8 | Diff (Post-Pre) | **1.33** | **1.62** | | |
| Anti-S IgG against B.1.351 | | | | | | | | |
| Pre | 38.0 | 14.0 | 138.5 | Pre | -0.81 | -0.69 | 1271 | 1.00 |
| Post | 1276.0 | 736.0 | 2844.0 | Post | 0.69 | 0.81 | 1277 | 1.00 |
| Fold change (Post-Pre/Pre) | 36.6 | 10.5 | 104.5 | Diff (Post-Pre) | **1.37** | **1.62** | | |
| Anti-S IgG against P.1 | | | | | | | | |
| Pre | 25.0 | 9.0 | 89.5 | Pre | -0.81 | -0.68 | 1048 | 1.00 |
| Post | 824.0 | 469.0 | 1917.0 | Post | 0.68 | 0.81 | 1052 | 1.00 |
| Fold change (Post-Pre/Pre) | 31.0 | 10.9 | 103.9 | Diff (Post-Pre) | **1.35** | **1.63** | | |

*ESS = Effective sample size

### Table S4: Bayesian ANOVA regression results: Serum neutralization

| Serum neutralization ($ID_{50}$) | | | | Bayesian ANOVA results | | | | |
|---|---|---|---|---|---|---|---|---|
| | | Interquartile range | | | 95% credible interval | | | |
| Variable | Median | Lower | Upper | Variable | Lower | Upper | ESS | $\hat{R}$ |
| **WT (D614G) SARS-CoV-2 - entire cohort** | | | | | | | | |
| Control | 1863.0 | 1321.0 | 3020.0 | Control | -0.16 | -0.02 | 852 | 1.00 |
| Pre | 714.0 | 213.0 | 1866.5 | Pre | -0.61 | -0.46 | 855 | 1.00 |
| Post | 10635.0 | 6312.0 | 17128.0 | Post | 0.56 | 0.67 | 856 | 1.00 |
| | | | | Post-Control | 0.59 | 0.81 | | |
| | | | | Pre-Control | -0.59 | -0.33 | | |
| Fold change (Post-Pre/Pre) | 12.5 | 5.2 | 39.6 | Diff (Post-Pre) | **1.05** | **1.27** | | |
| **WT (D614G) SARS-CoV-2 - randomly selected 20 participants** | | | | | | | | |
| Pre | 489.5 | 211.4 | 1753.3 | Pre | -0.78 | -0.49 | 1025 | 1.00 |
| Post | 12902.0 | 7674.0 | 17982.8 | Post | 0.49 | 0.78 | 1027 | 1.00 |
| Fold change (Post-Pre/Pre) | 23.4 | 9.5 | 37.0 | Diff (Post-Pre) | **0.98** | **1.56** | | |
| **B.1.1.7** | | | | | | | | |
| Pre | 398.4 | 100.0 | 1068.8 | Pre | -0.82 | -0.52 | 1171 | 1.00 |
| Post | 8261.5 | 7052.5 | 11410.8 | Post | 0.52 | 0.82 | 1159 | 1.00 |
| Fold change (Post-Pre/Pre) | 29.8 | 7.2 | 70.4 | Diff (Post-Pre) | **1.05** | **1.64** | | |
| **B.1.351** | | | | | | | | |
| Pre | 190.4 | 100.0 | 885.3 | Pre | -0.69 | -0.33 | 1069 | 1.00 |
| Post | 3297.5 | 1767.0 | 7220.0 | Post | 0.33 | 0.69 | 1050 | 1.00 |
| Fold change (Post-Pre/Pre) | 9.1 | 6.4 | 21.2 | Diff (Post-Pre) | **0.67** | **1.37** | | |
| **P.1** | | | | | | | | |
| Pre | 438.8 | 105.9 | 1364.0 | Pre | -0.73 | -0.39 | 1465 | 1.00 |
| Post | 7099.5 | 3940.3 | 11111.8 | Post | 0.39 | 0.73 | 1481 | 1.00 |
| Fold change (Post-Pre/Pre) | 10.5 | 5.7 | 36.8 | Diff (Post-Pre) | **0.78** | **1.46** | | |
| **WT(D614G) v. B.1.1.7 (Pre-vaccination)** | | | | | | | | |
| WT | 489.5 | 211.4 | 1753.3 | WT | -0.08 | 0.25 | 1011 | 1.01 |
| VOC | 398.4 | 100.0 | 1068.8 | VOC | -0.25 | 0.08 | 1002 | 1.01 |
| | | | | Diff (VOC-WT) | -0.50 | 0.16 | | |
| **WT(D614G) v. B.1.351 (Pre-vaccination)** | | | | | | | | |
| WT | 489.5 | 211.4 | 1753.3 | WT | -0.07 | 0.28 | 956 | 1.00 |
| VOC | 190.4 | 100.0 | 885.3 | VOC | -0.28 | 0.07 | 945 | 1.00 |
| | | | | Diff (VOC-WT) | -0.56 | 0.13 | | |
| **WT(D614G) v. P.1 (Pre-vaccination)** | | | | | | | | |
| WT | 489.5 | 211.4 | 1753.3 | WT | -0.11 | 0.21 | 465 | 1.01 |
| VOC | 438.8 | 105.9 | 1364.0 | VOC | -0.21 | 0.11 | 471 | 1.01 |
| | | | | Diff (VOC-WT) | -0.42 | 0.23 | | |

| | | | | | | | | |
|---|---|---|---|---|---|---|---|---|
| WT(D614G) v. B.1.1.7 (Post-vaccination) | | | | | | | | |
| WT | 12902.0 | 7674.0 | 17982.8 | WT | -0.04 | 0.14 | 688 | 1.01 |
| VOC | 8261.5 | 7052.5 | 11410.8 | VOC | -0.14 | 0.04 | 698 | 1.01 |
| | | | | Diff (VOC-WT) | -0.28 | 0.07 | | |
| WT(D614G) v. B.1.351 (Post-vaccination) | | | | | | | | |
| WT | 12902.0 | 7674.0 | 17982.8 | WT | 0.12 | 0.38 | 1065 | 1.00 |
| VOC | 3297.5 | 1767.0 | 7220.0 | VOC | -0.38 | -0.12 | 1076 | 1.01 |
| | | | | Diff (VOC-WT) | **-0.76** | **-0.23** | | |
| WT(D614G) v. P.1 (Post-vaccination) | | | | | | | | |
| WT | 12902.0 | 7674.0 | 17982.8 | WT | -0.01 | 0.24 | 976 | 1.00 |
| VOC | 7099.5 | 3940.3 | 11111.8 | VOC | -0.24 | 0.01 | 978 | 1.00 |
| | | | | Diff (VOC-WT) | -0.48 | 0.02 | | |
| B.1.1.7/WT ratio | | | | | | | | |
| Pre | 0.82 | 0.56 | 1.06 | Pre | -0.14 | 0.14 | 738 | 1.00 |
| Post | 0.73 | 0.58 | 1.02 | Post | -0.14 | 0.14 | 676 | 1.00 |
| | | | | Diff (Post-Pre) | -0.28 | 0.28 | | |
| B.1.351/WT ratio | | | | | | | | |
| Pre | 0.85 | 0.53 | 1.00 | Pre | 0.04 | 0.45 | 937 | 1.00 |
| Post | 0.36 | 0.20 | 0.44 | Post | -0.45 | -0.04 | 895 | 1.01 |
| | | | | Diff (Post-Pre) | **-0.90** | **-0.08** | | |
| P.1/WT ratio | | | | | | | | |
| Pre | 0.90 | 0.69 | 1.36 | Pre | -0.03 | 0.38 | 742 | 1.00 |
| Post | 0.59 | 0.44 | 0.80 | Post | -0.38 | 0.03 | 737 | 1.00 |
| | | | | Diff (Post-Pre) | -0.76 | 0.05 | | |

*****ESS = Effective sample size

**Table S5: Characteristics of study participants included in vaccine response - meta variables regression analysis**

| | No. of participants (%) |
|---|---|
| Total | 139 |
| Age (median = 50y. range = (22y. 80y) | |
| ≥45 y | 53 (38.1%) |
| 46-65 y | 63 (45.3%) |
| >65 y | 23 (16.5%) |
| Sex | |
| Female | 53 (38.1%) |
| Male | 86 (61.9%) |
| Severity | |
| Mild | 49 (35.3%) |
| Moderate | 62 (44.6%) |
| Severe | 13 (9.4%) |
| Critical | 15 (10.8%) |
| Time between symptom onset and vaccination (median = 9 months; range = (1 month. 15 months) | |
| ≥6 months | 47 (33.8%) |
| 7-12 months | 58 (41.7%) |
| >12 months | 34 (24.5%) |
| Comorbidities | |
| Cancer | 8 (5.8%) |
| Cardiovascular disease | 23 (16.5%) |
| Chronic respiratory disease | 19 (13.7%) |
| Diabetes Mellitus | 10 (7.2%) |
| Obesity | 25 (18.0%) |

**Table S6: Bayesian multilevel regression results: Post-vaccination neutralization titers v. participants' metadata**.

| | With B.1.1.7 | | | | Without B.1.1.7 | | | |
|---|---|---|---|---|---|---|---|---|
| | 95% credible interval | | | | 95% credible interval | | | |
| Variable | Lower | Upper | ESS* | $\hat{R}$ | Lower | Upper | ESS | $\hat{R}$ |
| Age | -0.29 | -0.05 | 5758 | 1.00 | -0.29 | -0.04 | 6872 | 1.00 |
| Sex | 0.04 | 0.32 | 4732 | 1.00 | 0.03 | 0.32 | 6387 | 1.00 |
| Severity | -0.02 | 0.17 | 4242 | 1.00 | -0.01 | 0.17 | 6681 | 1.00 |
| Time since symptom onset | -0.07 | 0.15 | 4148 | 1.00 | -0.09 | 0.13 | 6196 | 1.00 |
| Pre-vaccination neutralization | 0.16 | 0.45 | 2258 | 1.00 | 0.18 | 0.47 | 3359 | 1.00 |
| Cancer | -0.48 | 0.21 | 3667 | 1.00 | -0.48 | 0.19 | 7007 | 1.00 |
| Cardiovascular disease | -0.18 | 0.27 | 3959 | 1.00 | -0.19 | 0.27 | 5622 | 1.00 |
| Chronic respiratory disease | -0.32 | 0.12 | 1540 | 1.00 | -0.31 | 0.12 | 5602 | 1.00 |
| Diabetes Mellitus | -0.48 | 0.1 | 5077 | 1.00 | -0.49 | 0.08 | 7814 | 1.00 |
| Obesity | -0.14 | 0.23 | 6954 | 1.00 | -0.12 | 0.25 | 7814 | 1.00 |
| B.1.1.7 infection | -0.88 | 0.06 | 4479 | 1.00 | | | | |

*****ESS = Effective sample size

**Supplementary Methods**

To identify and estimate the effect size of different predictor variables on the observed pseudovirus SARS-CoV-2 neutralization data. we used a Bayesian hierarchical model that partially pooled effect size estimates across all study participants $l$. We assumed a linear correlation between the mean-centered predicted log neutralization values $\langle Y \rangle$ and predictor variables $X_i$:

$$\langle Y \rangle = \beta_{l.0} + \sum_i \beta_{l.i} X_i$$

where $\beta_{l.i}$ is the normalized effects of variable $i$ for participant $l$ and $\beta_{l.0}$ is the participant-specific intercept.

We assumed that the observed mean-centered and scaled neutralization values $Y$ follow a Student-T distribution about the predicted $\langle Y \rangle$ with error-term standard deviation $\sigma_Y$ with $\nu_Y$ degrees of freedom:

$$Y \sim T(\nu_Y . \langle Y \rangle . \sigma_Y)$$

We assumed that $\nu$ is exponentially distributed with a mean of 30 such that high prior probability was allocated over parameter values that describe the range from normal to heavy-tailed data under the Student-T distribution (Kruschke. 2011):

$$\nu \sim \mathrm{Exp}\left(\frac{1}{30}\right)$$

The intercepts $\beta_{l.0}$ were assumed to be normally distributed about a common mean intercept $\langle \beta_0 \rangle$ with standard deviation $\sigma_{\beta_0}$:

$$\beta_{l.0} \sim N\left(\langle \beta_0 \rangle . \sigma_{\beta_0}\right)$$

The participant-specific effect sizes $\beta_{l.i}$ of variable $i$ were assumed to be normally distributed about a common mean effect size $\langle \beta_i \rangle$ with a predictor-specific standard deviation $\sigma_{\beta_i}$:

$$\beta_{l.i} \sim N\left(\langle \beta_i \rangle . \sigma_{\beta_i}\right)$$

Weakly informative priors were placed on all standard deviation terms to constrain parameter inferences within biologically and mathematically plausible values (Gelman. 2006):

$$\sigma_Y \sim \text{Half-Normal}(0.1)$$
$$\sigma_{\beta_0} \sim \text{Half-Normal}(0.1)$$
$$\sigma_{\beta_i} \sim \text{Half-Normal}(0.1)$$

A weakly informative Gaussian prior was also placed for the mean intercept $\langle \beta_0 \rangle$ while a weakly informative Student-T prior was placed on the mean effect size $\langle \beta_i \rangle$ for each predictor $i$:

$$\langle \beta_0 \rangle \sim N(0.1)$$
$$\langle \beta_i \rangle \sim T(3.\,0.\,2.5)$$

We performed three different analyses with the aforementioned model correlating different predictors to mean-centered log neutralisation values (Table S1).

We also implemented a Bayesian hierarchical generalisation of the one-way ANOVA model to estimate the bounds on the effects of individual groups $j$ of a nominal predictor $i$ on an observed metric variable. We assumed that the predicted mean-centered metric variable $\langle Y_i \rangle$ is given by:

$$\langle Y_i \rangle = \beta_{i.0} + \sum_j \beta_{i.j} x_{i.j}$$

where $x_{i.j}$ is a Boolean variable denoting if an individual belongs to subgroup $j$ for the nominal predictor $i$.

We assumed that the observed metric data ($Y_i$) can be described by the Student-t distribution with $\nu$ degrees of freedom. the predicted location $\langle Y_i \rangle$ and heterogenous variances for individual groups $\sigma_{i.[j]}$:

$$Y \sim T\left(\nu . \langle Y_i \rangle . \sigma_{i.[j]}\right)$$

The intercept $\beta_{i.0}$ was again placed with a weakly informative Gaussian prior:

$$\beta_{i.0} \sim N(0.1)$$

We placed Student-T prior on the effect size $\beta_{i.j}$ for each subgroup $j$ of nominal predictor $i$ centered around zero. with weakly-informative gamma prior on $\nu_\beta$ degrees of freedom and positive-constrained half-normal prior on the standard deviation error-term $\sigma_\beta$:

$$\beta_{i.j} \sim T\left(\nu_\beta . 0 . \sigma_\beta\right)$$
$$\nu_\beta \sim \text{Gamma}(2.0.1)$$
$$\sigma_\beta \sim \text{Half-Normal}(0.1)$$

Following Kruschke (Kruschke. 2011). we assumed that $\nu$ is exponentially distributed with a mean of 30 such that high prior probability was allocated over parameter values that describe the range from normal to heavy-tailed data under the Student-T distribution:

$$\nu \sim \text{Exp}\left(\frac{1}{30}\right)$$

As mentioned earlier. we assumed a heterogenous scale term $\sigma_{i.j}$ for each subgroup $j$ that follows a gamma distribution with mode $\omega$ and standard deviation $\sigma_\sigma$. We placed vague gamma priors on both $\omega$ and $\sigma_\sigma$ that are broad on the scale of the data by estimating the shape ($k$) and scale ($\theta$) of the priors such that the mode and standard deviation equal to $\frac{\sigma_{Y_{obs}}}{2}$ and $2\sigma_{Y_{obs}}$ respectively:

$$\sigma_{i.j} \sim \text{Gamma}\left(k = 1 + \frac{\omega}{\theta} . \theta = \frac{2\sigma_\sigma^2}{\omega + \sqrt{\omega^2 + 4\sigma_\sigma^2}}\right)$$

$$\omega \text{ or } \sigma_\sigma \sim \text{Gamma}\left(k = 1 + \frac{\sigma_{Y_{obs}}}{2\theta} . \theta = \frac{2\left(2\sigma_{Y_{obs}}\right)^2}{\frac{\sigma_{Y_{obs}}}{2} + \sqrt{\left(\frac{\sigma_{Y_{obs}}}{2}\right)^2 + 4\left(2\sigma_{Y_{obs}}\right)^2}}\right)$$

We fitted all Bayesian models using Markov Chain Monte carlo (MCMC) with pymc3 (Salvatier et al.. 2016). implementing a no-u-turn sampler. Four MCMC chains were ran with at least 4000 burn-in steps and 2000 saved posterior samples. Convergence for all parameters were verified by checking trace plots. ensuring their $\hat{R}$ values were < 1.05 with sufficient effective sample size (>200).
